# Therapies for Long COVID in non-hospitalised individuals - from symptoms, patient-reported outcomes, and immunology to targeted therapies (The TLC Study): Study protocol

**DOI:** 10.1101/2021.12.20.21268098

**Authors:** Shamil Haroon, Krishnarajah Nirantharakumar, Sarah E Hughes, Anuradhaa Subramanian, Olalekan Lee Aiyegbusi, Elin Haf Davies, Puja Myles, Tim Williams, Grace M Turner, Joht Singh Chandan, Christel McMullan, Janet Lord, David Wraith, Kirsty McGee, Alastair Denniston, Tom Taverner, Louise Jackson, Elizabeth Sapey, Georgios Gkoutos, Krishna Gokhale, Edward Leggett, Clare Iles, Christopher Frost, Gary McNamara, Amy Bamford, Tom Marshall, Dawit Zemedikun, Gary Price, Steven Marwaha, Nikita Simms-Williams, Kirsty Brown, Anita Walker, Karen Jones, Karen Matthews, Jennifer Camaradou, Michael Saint-Cricq, Sumita Kumar, Yvonne Alder, David E. Stanton, Lisa Agyen, Megan Baber, Hannah Blaize, Melanie Calvert

## Abstract

**Introduction:** Individuals with COVID-19 frequently experience symptoms and impaired quality of life beyond 4-12 weeks, commonly referred to as Long COVID. Whether Long COVID is one or several distinct syndromes is unknown. Establishing the evidence base for appropriate therapies is needed. We aim to evaluate the symptom burden and underlying pathophysiology of Long COVID syndromes in non-hospitalised individuals and evaluate potential therapies.

**Methods and analysis:** A cohort of 4000 non-hospitalised individuals with a past COVID-19 diagnosis and 1000 matched controls will be selected from anonymised primary care records from the Clinical Practice Research Datalink (CPRD) and invited by their general practitioners to participate on a digital platform (Atom5™). Individuals will report symptoms, quality of life, work capability, and patient reported outcome measures. Data will be collected monthly for one year.

Statistical clustering methods will be used to identify distinct Long COVID symptom clusters. Individuals from the four most prevalent clusters and two control groups will be invited to participate in the BioWear sub-study which will further phenotype Long COVID symptom clusters by measurement of immunological parameters and actigraphy.

We will review existing evidence on interventions for post-viral syndromes and Long COVID to map and prioritise interventions for each newly characterised Long COVID syndrome. Recommendations will be made using the cumulated evidence in an expert consensus workshop. A virtual supportive intervention will be coproduced with patients and health service providers for future evaluation.

Individuals with lived experience of Long COVID will be involved throughout this programme through a patient and public involvement group.

**Ethics and dissemination:** Ethical approval was obtained from the Solihull Research Ethics Committee, West Midlands (21/WM/0203). The study is registered on the ISRCTN Registry (1567490). Research findings will be presented at international conferences, in peer-reviewed journals, to Long COVID patient support groups and to policymakers.

**Article Summary:** *Strengths and limitations of the study:*

- The study will generate a nationally representative cohort of individuals with Long COVID recruited from primary care.
- We will recruit controls matched on a wide range of demographic and clinical factors to assess differences in symptoms between people with Long COVID and similar individuals without a history of COVID-19.
- We will use a newly developed electronic patient reported outcome measure (Symptom Burden Questionnaire™) for Long COVID to comprehensively assess a wide range of symptoms highlighted by existing literature, patients, and clinicians.
- Immunological, proteomic, genetic, and wearable data captured in the study will allow deep phenotyping of Long COVID syndromes to help better target therapies.
- A limitation is that a significant proportion of non-hospitalised individuals affected by COVID-19 in the first wave of the pandemic will lack confirmatory testing and will be excluded from recruitment to the study.

## Introduction

Coronavirus Disease 2019 (COVID-19), caused by Severe Acute Respiratory Syndrome Coronavirus 2 (SARS CoV-2), is the most significant pandemic since the Spanish Influenza Pandemic of 1918. There have been over 240 million cases worldwide, including over 8.4 million cases in the UK.[1,2] In the UK, this has resulted in over 550,000 hospital admissions and 160,000 deaths.[3,4]

SARS CoV-2 enters human cells using the angiotensin II receptor, which is widely expressed throughout the body. COVID-19 is consequently a multisystem disorder affecting the lungs, heart, gut, kidneys, brain, and skin. A wide range of symptoms have been associated with COVID-19 including breathlessness, fatigue, fever, myalgia, “brain fog” and anosmia.[5]

Symptoms resolve within 12 weeks in most affected individuals. However, in up to 10% of individuals, symptoms of COVID-19 persist, sometimes in a relapsing remitting manner.[6] This can have significant impacts on physical and neurocognitive functioning, quality of life and work capability.[5] Those having symptoms lasting beyond 12 weeks are referred to as experiencing post-COVID-19 syndrome (NICE 2019) or more widely known as Long COVID.[7]

The cause of Long COVID is poorly understood. However, there is evidence from other post-viral syndromes that may be applicable to Long COVID. Evidence from existing studies on COVID-19 suggest a significant burden of pathophysiological insults and sequelae such as lung scarring, kidney injury, myocarditis and systemic inflammatory states that may promulgate longer term symptoms.[5] It is possible that autoimmune pathways may be triggered by COVID-19, leading to multi-system inflammatory damage.[8]

Current evidence suggests that 2.3-37% of individuals with COVID-19 experience symptoms and impaired quality of life and frequently report experiencing heterogeneous physical and psychological symptoms beyond 12 weeks (“Long COVID”) that can be debilitating.[9–11] People living with Long COVID have indicated that they are suffering with a range of symptoms, feel ‘abandoned’ and ‘dismissed’ by healthcare providers and receive limited or conflicting advice.[12] The aetiology, pathophysiology and treatments for Long COVID are not well understood, creating an unmet need for the growing number of affected individuals. Although efforts are being made to study Long COVID in hospitalised patients, there is a large unmet need among non-hospitalised individuals. Long COVID may comprise several distinct syndromes yet to be fully characterised.

Existing evidence on Long COVID is based on hospitalised cohorts and non-selected populations that are unlikely to be generalisable to the wider UK population of non-hospitalised individuals. A representative population-based cohort with well-matched controls, ideally derived from primary care, is needed to understand the burden of Long COVID, associated disability and impact on work capability. Current literature suggests the physical and psychological symptoms of Long COVID are highly diverse and may represent several distinct syndromes.[13] The characterisation of these syndromes using real-world data in combination with patient reported outcomes would aid health services to deliver appropriate interventions to meet these health needs.

The aetiology and risk factors for Long COVID also need further investigation as literature suggests a disproportionately higher prevalence of Long COVID among women, older adults and individuals with specific symptom clusters.[11,14,15] This may be due to differing immunological profiles of individuals who go on to develop Long COVID.[16,17] For example, older adults with COVID-19 show higher levels of senescent T cells.[18] As these are pro-inflammatory, more likely to be autoimmune and tissue-damaging, their persistence could mediate distinct symptoms in Long COVID among these individuals. Understanding the immunological basis of Long COVID would better enable clinicians to offer targeted therapies.[19]

There is an urgent need for evidence-based, accessible interventions, co-produced with patients with lived experience of COVID-19, to better support the growing number of non-hospitalised individuals with Long COVID. We propose to derive treatment recommendations drawing upon existing approaches to the management of post-viral syndromes in addition to a detailed understanding of Long COVID syndromes.

There is also a need to develop a trial platform that can link symptom, primary care, and hospital data with immunological profiling to evaluate interventions for Long COVID in non-hospitalised patients. The Recovery trial and PHOSP-COVID study have demonstrated the strength of platforms where several interventions can be rapidly evaluated in secondary care.[20,21] The PRINCIPLE study was set up for non-hospitalised patients in primary care with acute COVID-19.[22] However, such an infrastructure is still needed for affected individuals with Long COVID in the wider community, and studies such as STIMULATE-ICP are being set up to help address this.[23]

The TLC Study will directly address the patient needs highlighted in the NIHR Evidence Report, by the LongCovidSOS campaign, our patient and public involvement group and patient partners, who have co-designed this study.[24,25] Our research will characterise the symptoms, health impacts and underlying pathophysiology of Long COVID, define its component syndromes in non-hospitalised individuals, and provide resources to support symptom management and nurse-led support for those with the severest symptoms. We will use our findings to co-produce with patients a targeted intervention for Long COVID, tailored to individual patient needs.

## Aims

The aim of the study is to evaluate the symptom burden and underlying pathophysiology of Long COVID syndromes in non-hospitalised individuals and the impact on quality of life and work capability. We also aim to identify potential therapies and co-produce a remotely delivered intervention for supporting individuals with Long COVID syndromes. These findings will be used to inform a feasibility study to evaluate the intervention produced during this project, and to plan future evaluations of supportive and pharmacological interventions for treating Long COVID. The methods for therapy identification, intervention development and feasibility testing will be reported in separate linked protocols.

The key objectives are:

1. To establish a representative population-based cohort of non-hospitalised individuals with COVID-19 and prolonged symptoms (“Long COVID”).
2. To validate a new patient-reported outcome (PRO) measure of symptom burden in individuals with Long COVID- the Symptom Burden Questionnaire ™ for Long COVID.
3. To collect validated PROs including symptoms, quality of life and work capability data longitudinally from the representative cohort of non-hospitalised individuals with Long COVID.
4. To identify distinct symptom clusters (“syndromes”) in individuals with Long COVID and compare these Long COVID syndromes to known post-viral syndromes.
5. To measure immunological, proteomic and miRNA biomarkers to determine the underlying pathogenesis associated with Long COVID syndromes. To capture physical health measures via a wearable device to determine the physical exertions (heart rate intensity and movement) and sleep patterns observed in Long COVID syndromes.
6. To make recommendations on pharmacological and supportive therapies that: i) should be implemented without need for further evaluation; ii) should be evaluated in a clinical trial, and iii) should not be recommended for treating long COVID based on the available evidence.
7. Co-produce a supportive, remotely delivered intervention for individuals with Long COVID, to address the symptoms associated with each Long COVID syndrome.
8. To evaluate patient and public involvement in the study.

## Methods and analysis

### Study design

We will undertake a population-based cohort study of non-hospitalised individuals with Long COVID (cases) and matched controls recruited from general practices in England that contribute data to the Clinical Practice Research Datalink (CPRD) Aurum database (Figure 1).[26] CPRD is a real-world research service supporting retrospective and prospective public health and clinical studies. CPRD is jointly sponsored by the Medicines and Healthcare products Regulatory Agency and the National Institute for Health Research (NIHR), as part of the Department of Health and Social Care. CPRD collects anonymised patient data from a network of GP practices across the UK. Primary care data are linked to a range of other health related data to provide a longitudinal, representative UK population health dataset. The data in CPRD Aurum encompass 40 million research acceptable patients, including 13 million currently registered patients.

**Figure 1.**
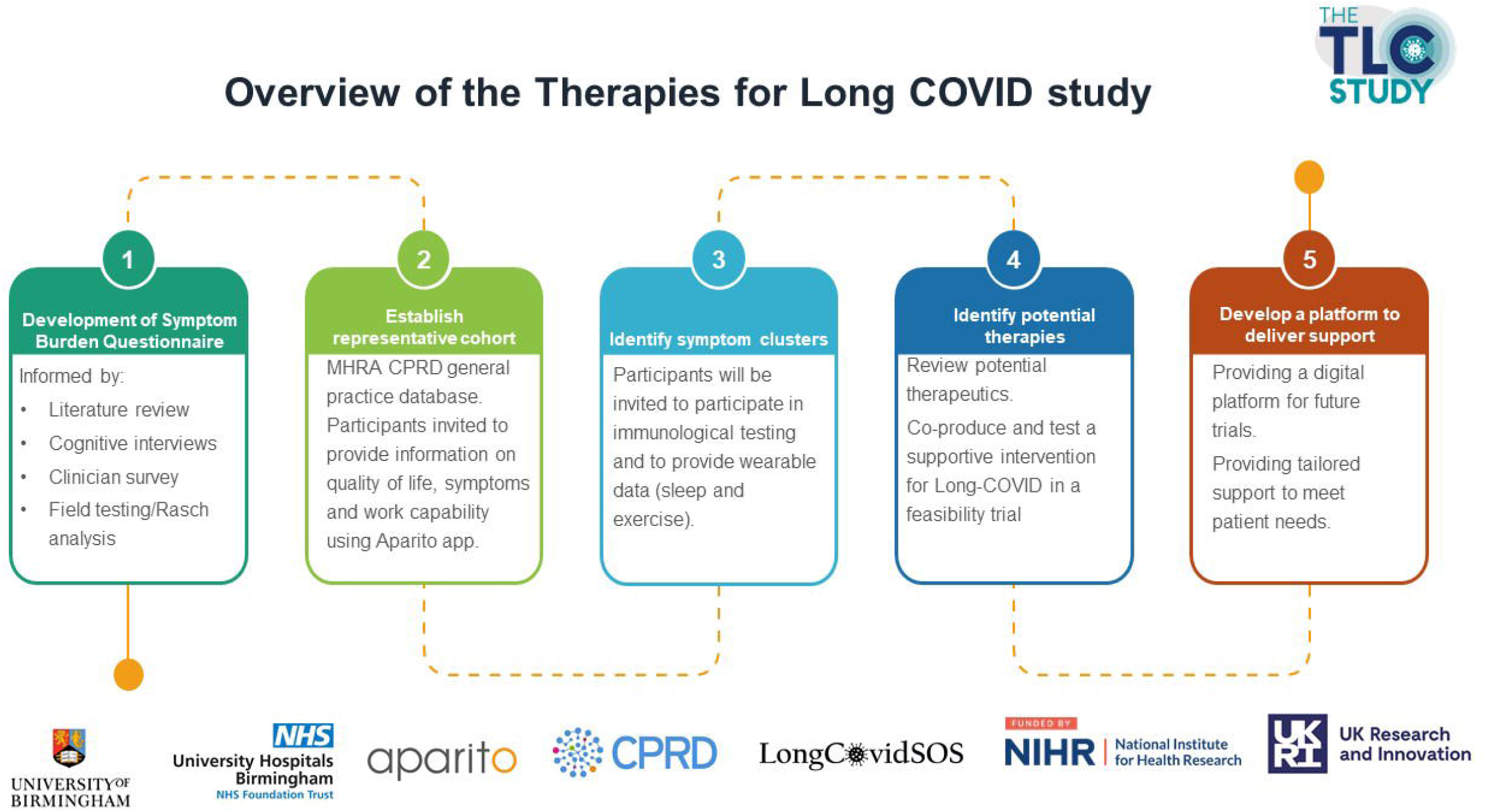
Project overview

Eligible cases and controls will be invited to participate in the study and to report their symptoms, quality of life and work capability monthly for 12 months on a digital platform called Atom5™ produced by the medical technology company Aparito Ltd. Cluster analysis methods will be used to define symptom clusters, which will be further described using data from linked GP records, hospital episode statistics and Office for National Statistics (ONS) mortality records.

A subgroup of participants sampled from each symptom cluster and two matched control groups (those with no history of COVID-19, and those with a history of COVID-19 but who did not develop Long COVID) will be invited to provide blood and saliva samples to measure their immune function, and proteomic and genomic profile (BioWear sub-study). These participants will also be provided a Garmin wearable device to monitor heart rate, oxygen saturation, step count, sleep quality, and post-exertional effects.

### Population and setting

Eligible practices and participants will be identified from the CPRD Aurum database[26] following data extraction using the Data Extraction for Epidemiological Research (DExTER) tool.[27] Practices to enrol in the cohort study will be selected using probability weighted sampling to ensure adequate geographic representation and oversampling of practices with larger numbers of patients with a history of COVID-19, low socioeconomic status, and higher proportions from Black and Ethnic Minority groups, while also preserving the balance of urban versus rural practices. Eligible patients from those practices will be flagged, with oversampling of cases from lower socioeconomic groups, ethnic minorities, and those who received a SARS CoV-2 RT-PCR positive result in 2021.

Participants will need to be actively registered on 31^st^ January 2020 and have at least 12 months of data prior to 31^st^ January 2020. Cases will be defined as those aged 18 years and older with a record of a positive SARS CoV-2 RT-PCR result (index date) and no record of hospitalization within 28 days of the diagnosis. Controls will be defined as individuals without a record of a positive SARS CoV-2 RT-PCR result or suspected or confirmed COVID-19 diagnosis at any time point from 31^st^ January 2020. Each case will be matched to four controls using propensity scores derived from demographic factors (age, sex, ethnic group, and socioeconomic status) and a comprehensive list of comorbidities. Controls will only be eligible if they have not been hospitalized within 28 days of the matched index date.

### Participant recruitment

Participating GP practices will be informed of eligible participants and asked to check their appropriateness for study invitation through the CPRD’s Interventional Research Services Platform.

Those verified as meeting the eligibility criteria and assessed as appropriate for study invitation by their general practitioner will be sent an invitation letter and patient information sheet, which will provide instructions on how to download the Atom5™ app or access the web portal and a unique onboarding code to register for the study. The study consent form will be hosted on Atom5™, which will include additional eligibility checks to ensure potential participants meet the study criteria. Participants unable to directly access Atom5™, for example due to inadequate technology access, will also be offered a telephone interview to complete the questionnaires with a research nurse. Language translators will also be provided when required.

### Patient reported outcomes (PROs)

Participants enrolled on Atom5™ will be asked to complete a novel electronic patient reported outcome (e-PRO) called the Symptom Burden Questionnaire™ for Long COVID (SBQ-LC™). This includes a comprehensive list of symptoms that were identified from a systematic review of symptoms associated with Long COVID,[5] discussions with Long COVID patients and clinicians, and field testing. Participants will also be asked to complete the Functional Assessment of Chronic Illness Therapy-Fatigue (FACIT-F), Patient Health Questionnaire-2 (PHQ-2), Generalised Anxiety Disorder-2 (GAD-2), Abbreviated PTSD Checklist (PCL-2), modified MRC Dyspnoea Scale,COVID-19 core outcome measure for recovery and Euroqol EQ-5D-5L and answer questions about work capability. Participants will be notified to complete these measures monthly for a total of 12 months.

### Defining symptom clusters

The symptom data captured at baseline in Atom5™ in addition to clinical data from primary care records will be used to define symptom clusters. We will employ clustering algorithms that have been shown to work well in medical settings, with binary and mixed variables: (1) complete linkage hierarchical clustering; (2) probabilistic c-means cluster analysis; and (3) density peaked clustering analysis. We will evaluate algorithm performance and the optimal number of clusters (e.g., by internal validation against a holdout dataset using metrics such as Gap statistic and silhouette index).[28,29]

The demographic and clinical characteristics of individuals in each symptom cluster will be described using data from Atom5™ and linked GP records in CPRD Aurum. This will include age, sex, ethnic group, socioeconomic status, smoking status, body mass index, SARS CoV-2 immunisation status, and comorbidities. Clinical outcomes for each of these clusters will be described using linked Hospital Episode Statistics and ONS mortality data.

### BioWear sub-study

50 participants sampled from the four most prevalent symptom clusters, 50 from the control group without a history of COVID-19, and 50 with a history of COVID-19 but without Long COVID, will be recruited to provide blood and saliva samples. These will be used to measure immune function (inflammatory markers, T-cell function, and autoantibodies), proteomics, and genomic profile. These participants will also be provided a Garmin Vivosmart 4 device to measure heart rate, oxygen saturation, step count and sleep quality at baseline, 6 months, and 12 months. Notifications will be sent once a month to undertake a 40-step test once a week for each week of device use, to assess for post-exertional effects of Long COVID. Together these data will allow for detailed immunological and digital phenotyping of the newly described symptom clusters.

### Sample size

We aim to recruit 4000 individuals with a history of COVID-19, with a minimum sample of 2000 (Figure 2). We also aim to recruit 1000 matched controls, with a minimum of 500. Reliable detection of clustered data via well-chosen combinations of these methods has been shown to require a minimum of 20-30 observations per subgroup provided good cluster separation exists (effect size Δ=4 or over).[30] We expect that with 500 patients in the Long Covid cohort we will have power above 80% to detect well-separated clusters comprising at least 5% of the cohort for K=4 to K=6 clusters.

**Figure 2.**
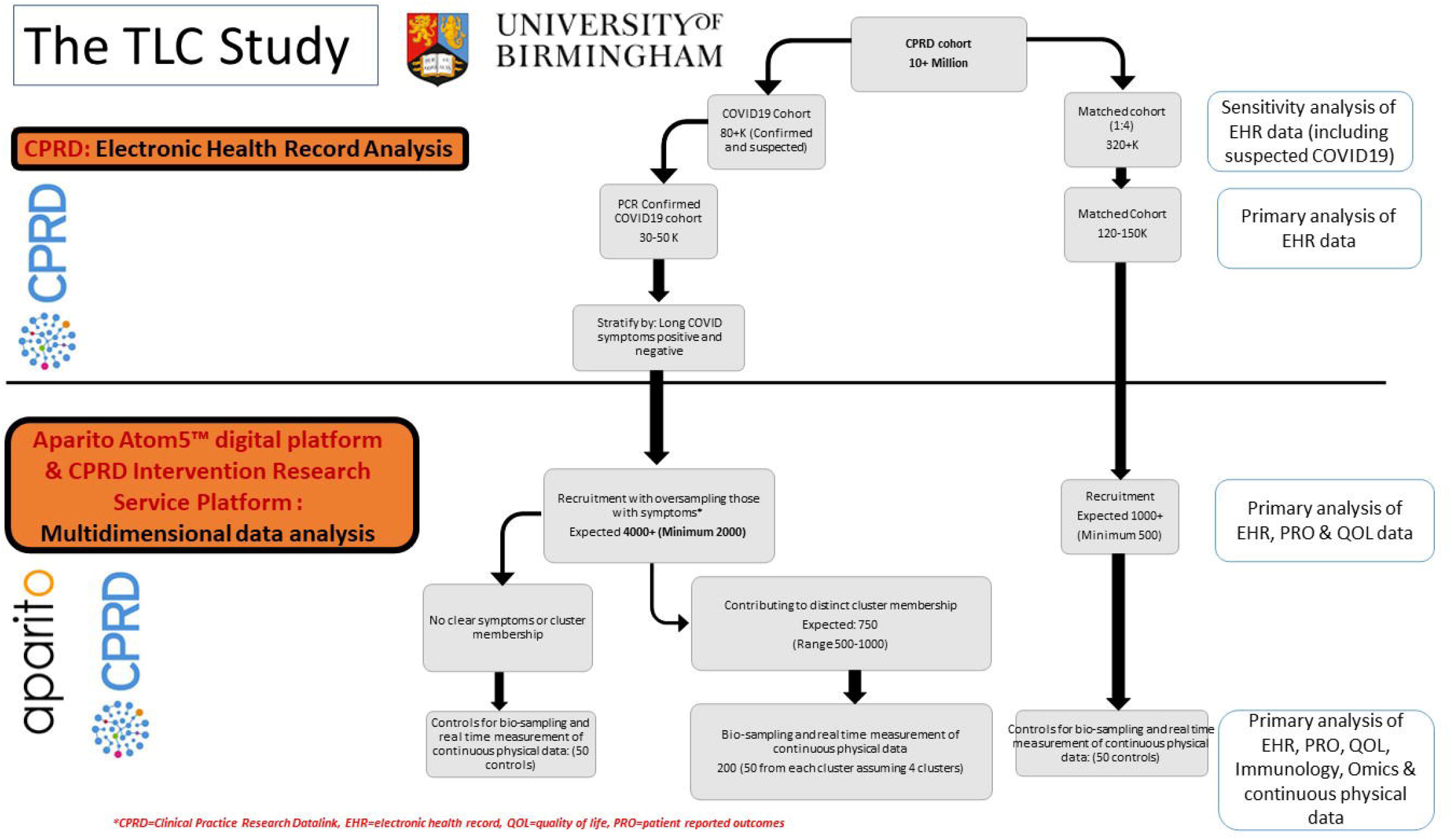
Study flow chart

### Analysis plan

Unsupervised exploratory clustering techniques will be employed to identify distinct Long COVID symptom clusters. We will first perform dimension reduction using one of: multi-dimensional scaling (MDS), t-stochastic neighbour embedding (t-SNE) and uniform manifold approximation and projection (UMAP). This first step will reduce dimensionality of the global data set while retaining local distances between individuals.

Following the dimensionality reduction step, we will employ clustering algorithms that have been shown to work well in medical settings:

1. k-means clustering, which locates the stable set of centroids of a predetermined number of k clusters.
2. Fuzzy c-means cluster analysis, a version of k-means clustering which allows data points to have partial cluster membership.
3. Hierarchical agglomerative clustering, which joins observations in a recursive fashion satisfying some linkage criterion until k clusters have been generated. We will use the Ward linkage, which joins observations that minimise the variance of merged clusters.
4. Gaussian finite mixture models, a model-based approach to clustering which associate each data point with a multivariate (ellipsoidal) Gaussian distribution.
5. DBSCAN/HDBSCAN, algorithms, which identify clusters of dense observations among unassigned lower-density observations.

The optimal number of clusters for all methods (except DBSCAN) will be determined using model selection methods and internal cross validation. Cluster membership goodness of fit will be evaluated by: (1) the Jaccard similarities (intersection size divided by union size) of the original clusters to the most similar clusters in the resampled data, and (2) the average silhouette score, which will vary between +1 for an observation aligned with its cluster centre and -1 for an observation aligned with the centre of another cluster. We will determine mean bootstrapped Jaccard and silhouette indices to assess cluster stability. The plot of within-groups sum of squares vs K will be checked graphically for optimal K (elbow method) and a minimum BIC.

### Other planned work

#### Epidemiology of Long COVID in primary care

We will undertake an epidemiological analysis of Long COVID symptoms and clinical outcomes using data from GP records in CPRD Aurum.[31] The prevalence of these symptoms will be compared to those in matched controls. Risk factors associated with the development of long-term symptoms in those with a history of COVID-19 will be described. Cluster analyses will be performed to identify distinct Long COVID phenotypes and their risk of hospital admission and mortality will be assessed.

#### Impact of Long COVID on healthcare utilisation and costs

The utilisation of a range of healthcare services (e.g., GP consultations, pharmacotherapy, secondary care referrals, and hospital admissions) within 12 months from the index date will be identified and enumerated for both cases and control groups, with UK unit costs (2021 values) assigned to each component.

To estimate the costs associated with primary care resource use we will measure different types of GP consultations (surgery visits, home visits, and telephone consultations). Unit costs (per visit) for each type of consultation will be derived from the Unit Costs of Health and Social Care (2020).[32] As it is not feasible to cost all medications prescribed for both the cases and control groups during the study period, we will focus only on a broad definition of relevant medications which could include respiratory (e.g., inhaled short acting beta-2 agonists) and cardiovascular (e.g., aspirin) therapies. The costs of prescribed medications will be obtained from NHS Electronic Drug Tariffs, BNF, or Prescription Cost Analysis. Costs associated with hospital admissions will be derived from linked HES Admitted Patient Care (APC) data. Other secondary care costs will be estimated based on secondary care referrals from primary care, which will include outpatient and accident and emergency referrals. The average costs per episode for each type of resource use will be obtained using NHS Reference Costs.[33]

The annual costs of healthcare utilisation for each different area of resource use will then be estimated as a product of the quantities of the resource per year and the attached unit costs. The total costs of all healthcare resource utilisation for each patient will be aggregated to make up the total direct costs of healthcare resource utilisation. Multivariable regression models will be used to estimate the incremental difference in the total all-cause healthcare costs between cases and controls controlling for demographic, clinical, and other confounding factors.

#### Impact of Long COVID on work capability and economic losses

To understand the broader impacts of long COVID on individuals and families, it is necessary to analyse work capability and economic losses. We will use lost productivity (absenteeism) as a proxy for indirect costs incurred since that is the most feasible cost component that can be captured in our data. Medically related absenteeism days will be estimated by identifying patients who reported work absence/changes in working hours using questions captured within Atom5™, and their linked sick notes record in CPRD. The associated costs will be calculated by applying average national earnings or based on individual employee wage information where available.

The human capital approach which is the most commonly used method for estimating the indirect costs of illnesses will be adopted.[34,35] This method equates the costs of illnesses to the losses of future total income that patients could have earned had they remained healthy, revealing the opportunity costs associated with long Covid in this case. Indirect costs incurred during the study period will be compared between the case and control groups using Wilcoxon signed-rank tests. Multivariable regression models will assess incremental cost differences between cases and controls, adjusting for confounding factors.

#### Post-viral syndromes: a systematic review of symptoms, health impacts, treatments, and their implications for Long COVID

This systematic review will narratively and quantitatively summarise the evidence on the prevalence of symptoms and health impacts (clinical complications and impacts on quality of life and work capability) associated with previous post-viral syndromes (e.g. post-viral effects of the Middle East Respiratory Syndrome). The review will also summarise the evidence from randomised controlled trials and observational studies on non-pharmacological treatments for post-viral syndromes including Long COVID. A review protocol will be published separately.

#### Survey of non-prescribed medication, supplements, remedies, and other therapies for Long COVID

A cross-sectional electronic survey will be undertaken to capture the non-prescribed medicines, supplements, remedies and other therapies used by individuals with long COVID to manage their symptoms, and what influenced their decision making. The aim is to identify self-prescribed therapies that are potentially harmful and pose clinical risks. This survey was suggested by members of the patient and public involvement group who reported their awareness of individuals self-medicating with a range of alternative therapies and highlighted the importance of capturing data on these self-management behaviours.

#### Consensus and co-production workshops

A series of expert consensus workshops will be convened to make recommendations on non-pharmacological therapies to be used in each Long COVID syndrome, to be evaluated in clinical trials, and to be recommended against, based on the cumulated evidence from the rapid systematic review and primary findings from WP2. The consensus workshop will follow the James Lind Alliance final priority setting method, based on Nominal Group Technique.[36] In addition, decision making will be guided by the APEASE criteria.[37] Particular consideration will be given to non-pharmacological therapies (e.g., physiotherapy, breathing techniques, sleep hygiene) that can be delivered digitally and at scale.

Subsequently, intervention co-production workshops will be held to determine which of the agreed interventions are suitable for or could be adapted for remote delivery. Person-centred design principles will be applied to co-produce a supportive, remotely delivered intervention, guided by the “building blocks of co-production” framework.[38]

Expert consensus and co-production workshops will be attended by people who have experienced Long COVID and their family members, friends, or carers; healthcare professionals; experts in symptoms and potential treatments relevant to Long COVID; regulators; and policy makers.

#### Feasibility study

A feasibility study will be undertaken on the newly coproduced intervention. This will assess the feasibility of participant recruitment, randomisation of eligible participants to the intervention and control groups, capture of intervention process measures, and collecting outcome and resource measures. The results of the feasibility study will help to determine whether to progress to a fully powered randomised controlled trial.

#### De-centralised trial platform

An important longer-term aim of the TLC Study is to develop a decentralised trial platform for evaluating therapies for Long COVID. This platform will be able to flag eligible participants using GP record data in CPRD Aurum, recruit participants using the CPRD Intervention Research Service Platform, obtain e-consent and baseline data on Atom5™, and obtain outcome data from a combination of Atom5™ for PROs, and clinical outcomes from GP records and linked hospital episode statistics and ONS mortality data.

#### Evaluation of PPI

Throughout the project, we will co-evaluate patient and public involvement (PPI) activities with the PPI members and obtain their views on how empowered they felt to meaningfully contribute to the study. The Guidance for Reporting Involvement of Patients and the Public (GRIPP) 2 checklist will be used for the reporting and evaluation of PPI for the TLC project.[39] Feedback on PPI activity outcomes will be shared with the PPI group.

## Patient and public involvement

A PPI group comprising of up to 15 individuals has been established following the NIHR Include guidance, which includes diverse ethnic and socioeconomic representation.[40] All study plans have been informed through discussions with the group and through broader discussions with national patient support groups (LongCovidSOS, Long Covid Scotland, Long Covid Wales, and Long Covid Support). This has significantly contributed to the development of the SBQ-LC™ and informed many aspects of the TLC study more broadly. Research findings will be discussed regularly with the group and revised based on their input accordingly. The PPI group will also inform the development of and contribute to dissemination plans, coauthor publications, and prepare lay friendly materials to disseminate research findings to patients and the public. Group members will be reimbursed for their time and any expenses incurred according to the UK standards for public involvement.[41]

## Ethics and dissemination

Ethical approval for the study was provided by the Solihull Research Ethics Committee (21/WM/0203).

## Data Availability

Data for this project are not currently available for access outside the research team. Each of the datasets generated within the project may be shared when finalised but this will require an application to the data controllers. The data may then be released to a specific research team for a specific project dependent on the independent approvals being in place.

